# Rapid vigilance and episodic memory decrements in COVID-19 survivors

**DOI:** 10.1101/2021.07.06.21260040

**Authors:** Sijia Zhao, Kengo Shibata, Peter J. Hellyer, William Trender, Sanjay Manohar, Adam Hampshire, Masud Husain

**Author notes:** Corresponding Author: Sijia Zhao, Department of Experimental Psychology, University of Oxford, Oxford OX1 3PH, UK. Abbreviations: COVID-19 = Coronavirus disease 2019 caused by the Coronavirus SARS- CoV-2; NFI = Neurological Fatigue Index; CFQ = Cognitive Failures Questionnaire; BFI-S = Short Big Five Inventory; GRIT-S = Short Grit Scale; AMI = Apathy Motivation Index; HADS = Hospital Anxiety and Depression Scale; SD = Standard Deviantion; GLM = Generalised linear model; BF = Bayesian Factor; PCA = Principal Component Analysis; LMM = Linear Mixed-effect Model; SES = socioeconomic status.

## Abstract

Recent studies indicate that COVID-19 infection can lead to serious neurological consequences in a small percentage of individuals. However, in the months following acute illness, many more suffer from fatigue, low motivation, disturbed mood, poor sleep and cognitive symptoms, colloquially referred to as ‘brain fog’. But what about individuals who had asymptomatic to moderate COVID-19 and report no concerns after recovering from COVID-19? Here we examined a wide range of cognitive functions critical for daily life (including sustained attention, memory, motor control, planning, semantic reasoning, mental rotation and spatial-visual attention) in people who had previously suffered from COVID-19 but were not significantly different from a control group on self-reported fatigue, forgetfulness, sleep abnormality, motivation, depression, anxiety and personality profile. Reassuringly, COVID-19 survivors performed well in most abilities tested, including working memory, executive function, planning and mental rotation. However, they displayed significantly worse episodic memory (up to 6 months post-infection) and greater decline in vigilance with time on task (for up to 9 months). Overall, the results show that specific chronic cognitive changes following COVID-19 are evident on objective testing even amongst those who do not report a greater symptom burden. Importantly, in the sample tested here, these were not significantly different from normal after six-nine months, demonstrating evidence of recovery over time.

## Introduction

People who survive COVID-19 infection present a significantly higher risk of major neurological and psychiatric conditions, particularly if they were hospitalised.^1–3^ These include acute cerebrovascular events such as ischaemic stroke and intracerebral haemorrhage.^4^ In addition to severe neurological conditions, there can also be more chronic, longer-term consequences such as fatigue, low motivation, disturbed mood and poor sleep—all commonly reported symptoms amongst survivors, the so-called ‘long-COVID’ (see recent review^5^).

While many studies employ questionnaires reliant on patients’ subjective self-reports, recent investigations have begun to document a wide range of post-COVID-19 cognitive deficits using objective cognitive testing of inpatients. Particular impairments were found in *attention*^6–12^, *memory*^6–8, 10–13^ and *executive functions.*^10–12, 14–16^ More recently, using ^18^F-FDG PET, it has been demonstrated that in the most severely affected patients, the degree of cognitive impairment was accompanied by frontoparietal hypometabolism.^10, 11^

Understandably, most of these small-scale reports have, to date, predominantly focused on symptomatic, hospitalised patients (however see^17^). But what about individuals who have not been hospitalised and do not report any ongoing symptoms after recovery? Do they suffer cognitive deficits that they are not aware of? Here, we examined people who had previously contracted COVID-19 but were not significantly different from a control group with respect to self-reported fatigue, forgetfulness, sleep abnormality, motivation, depression, anxiety or personality profile. In total, 135 participants were tested on a series of twelve on-line cognitive tasks encompassing a wide range of cognitive functions critical for daily life, including the ability to sustain attention, memory, motor control, speed of response, planning, semantic reasoning, mental rotation, and spatial-visual attention. In all of these functions, COVID-19 survivors showed the same initial baseline performance. However, after only two minutes on an attentionally demanding task, there was a significantly larger decline in perceptual sensitivity, despite reporting the same levels of fatigue compared to healthy controls. Subsequent testing also revealed a significantly larger episodic memory decrement. These results demonstrated that chronic cognitive reductions following COVID-19 are evident upon objective testing even in people who do not report long-COVID symptoms.

## Materials and methods

### Participants

155 participants (59 female, mean age 28.6 years (SD 9.7)) were recruited from the Prolific online recruitment platform (www.prolific.co). All participants were naïve about the aim of this study; the study was advertised as “a brain game” testing how well people could perform. 64 people had contracted COVID-19, while 91 reported that they had not. Although none had received treatment in an intensive care unit or were seeking post-COVID care at the time the study was conducted, we excluded three participants who had been hospitalised for COVID-19, seven participants who had severe COVID-19 symptoms (i.e., their COVID-19 symptoms largely reduced their ability to carry-out day-to-day activities) and two participants who had severe long-COVID symptoms. In addition, a follow-up survey showed that eight control participants later discovered that they contracted COVID-19 before via antigen detection test; although we did not find any difference in behavioural performance between them the rest of the control group, they were also excluded from further analysis.

In all, 136 participants were included in further analysis (COVID n=53, Control n=83, see **Fig 1** for study population flow and **Table 1** for demographics). There was no significant group difference in gender (*χ*^2^(1,N=54) = 0.5, p=0.5) or age (t(134)=-0.6, *p*=0.5, BF=4.4).

**Figure 1.**
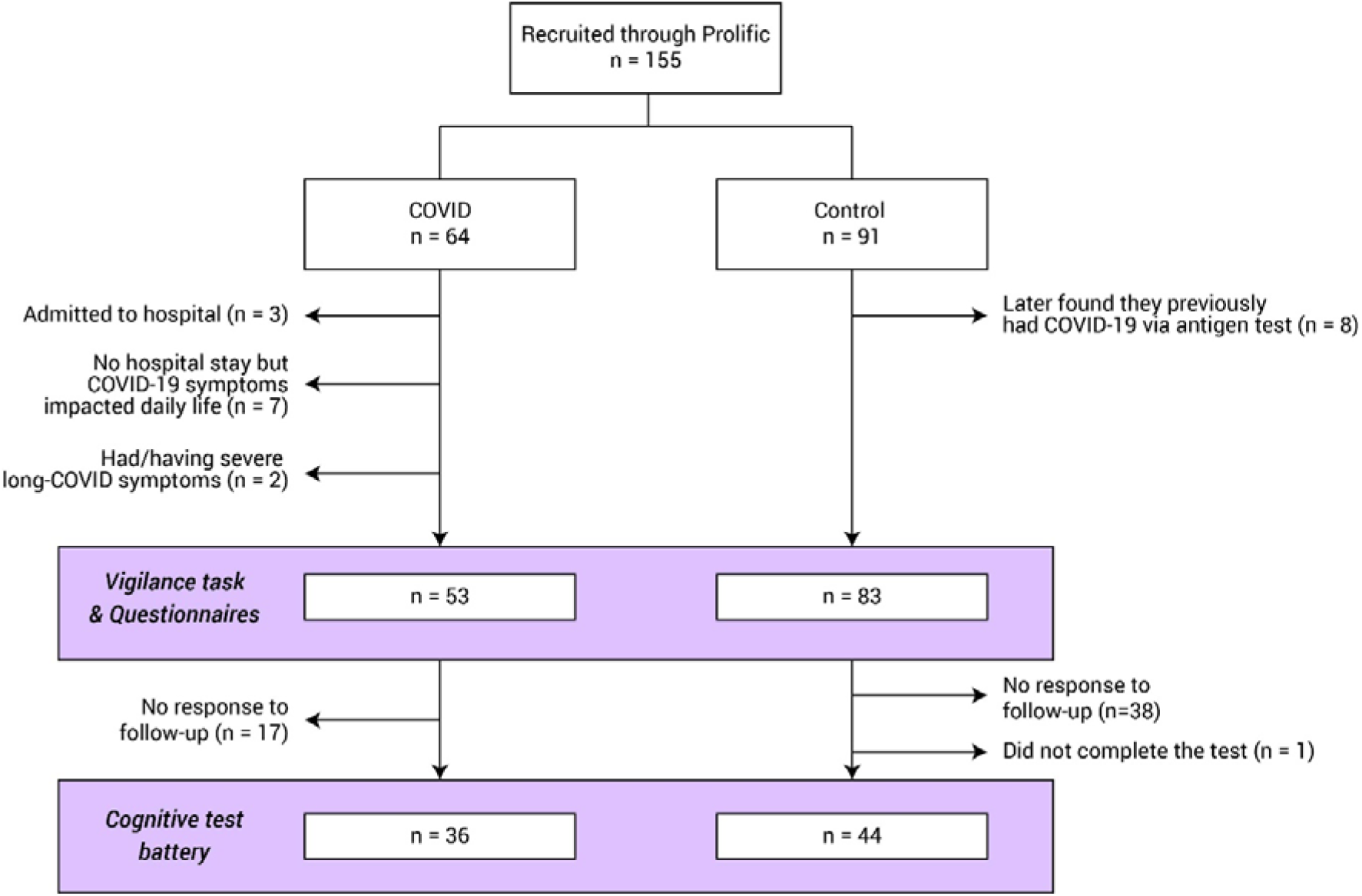
Study population flow chart. Number of participants eligible for each experimental session.

**Table 1.**
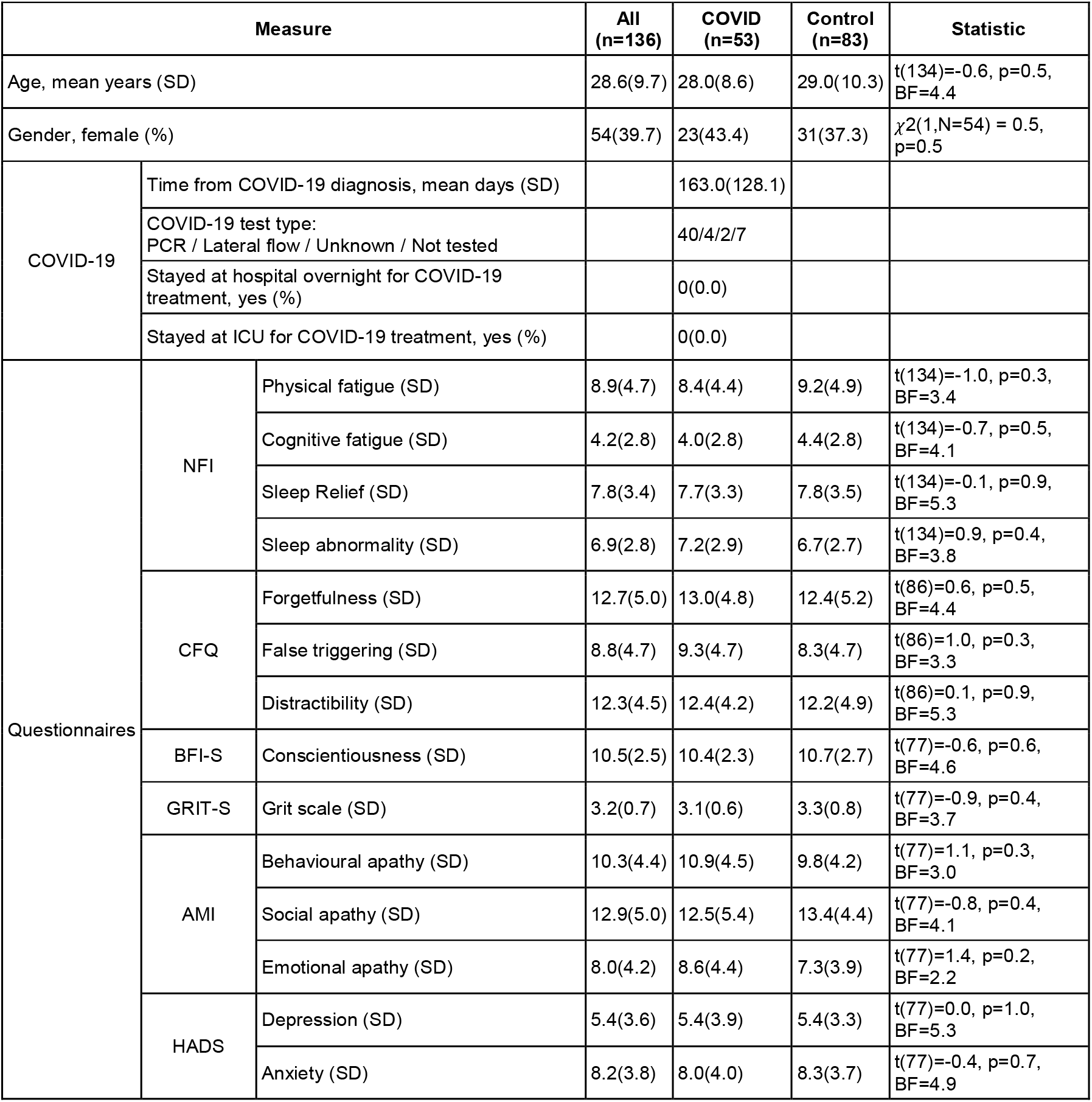
Self-reported participant demographics and questionnaire-derived measures. T- and χ^2^ tests were used to assess between-group differences, with Bayes Factor (BF) reported. The questionnaires included are Neurological Fatigue Index (NFI), Cognitive Failures Questionnaire (CFQ), Short Big Five Inventory (BFI-S), Short Grit Scale (GRIT-S), Apathy Motivation Index (AMI), and the Hospital Anxiety and Depression Scale (HADS). For all the questionnaire-derived indices, the mean score is shown with one standard deviation (SD) in the bracket.

All participants were required to complete the experiments on the Chrome browser on a desktop with a keyboard and mouse.

To compare the episodic memory deficit observed in the COVID group (see below), we ran the same memory test on healthy elderly people (>50 years old). 64 participants, who reported no neurological conditions, were recruited from the local community and have been involved in the lab’s previous studies. All participants provided written informed consent and the study was approved by the University of Oxford ethics committee. Participants were initially contacted by emailed, followed up by calls and completed the task on their own computer devices. 53 participants completed the task and one participant was excluded due to COVID-19 history. The results of 52 participants (30 females, mean age 67.4 years (SD 7.2)) are reported below.

### Questionnaires

The questionnaires included were:

**•Fatigue and sleep:** Neurological Fatigue Index (NFI), which has been used to assess fatigue in persons with multiple sclerosis.^18^ Our motivation for including this index was driven by the fact that it probes the interactions between sleep and fatigue. Specifically, it asks questions related to abnormal nocturnal sleep, sleepiness, and the need for diurnal sleep/rest.
**•Distractibility and forgetfulness:** Cognitive Failures Questionnaire (CFQ)^19^, a 25-item questionnaire about minor mistakes in daily life over the last two weeks. E.g.: “Do you find you forget appointments?” “Do you fail to notice signposts on the road?” “Do you find you forget which way to turn on a road you know well but rarely use?”.
**•Personality:** A short 15-item Big Five Inventory (BFI-S)^20^ and the Short Grit Scale (GRIT-S)^21^. One of BFI-S components—conscientiousness—provides information about the conscientiousness personality trait, describing an individual’s perseverance of effort combined with passions for a particular goal. To ensure that this is well-captured, we also included GRIT-S: this estimates the same trait but with different questions. We indeed found that these two scales were strongly positively correlated (Spearman’s rho = 0.65, *p*<0.0001).
**•Motivation:** Apathy Motivation Index (AMI), an 18-item questionnaire, sub-divided into three subscales of apathy: emotional, behavioural and social apathy.^22^
**•Mood:** Hospital Anxiety and Depression Scale (HADS), a 14-item questionnaire, sub- divided into depression and anxiety.^23^

All 155 participants completed the NFI questionnaire, of which 103 also completed the CFQ. 93 of these 103 participants completed the full set of questionnaires which were NFI, CFQ, BFI-S, GRIT-S, AMI, and HADS.

### Vigilance test

All participants were first tested on a version of an established, sustained visual attention task^24^ adapted into a modern online version hosted on the Pavlovia platform (pavlovia.org). An online demo is available at https://run.pavlovia.org/sijiazhao/vigilance_english_demo (please open it with the Chrome internet browser on a desktop computer). This task is designed to assess the performance decrement during sustained visual attention. A single digit (0-9) was presented at the centre of the screen for 50ms every second (**Fig 2** *Vigilance*). Participants were instructed to press the spacebar on their keyboard as soon as they saw “0” (the target, presented randomly with a probability of 25%); no response was required for other digits. A semi-transparent jittered checkerboard pattern masked the digits, with the level of difficulty manipulated by adjusting the opacity of the mask. Pilot testing allowed us to obtain an accuracy level of ∼80% in the first minute of the experiment. The practice phase consisted of 90 trials, equivalent to 90 seconds. The difficulty was gradually increased and feedback was provided after each trial. The first 12 practice trials were highly visible stimuli and participants were required to get 100% accuracy to proceed. Subsequent non-practice trials and blocks contained no feedback. In total, each participant completed 540 trials, divided into 9 blocks, each containing 60 trials and lasting one minute.

**Figure 2.**
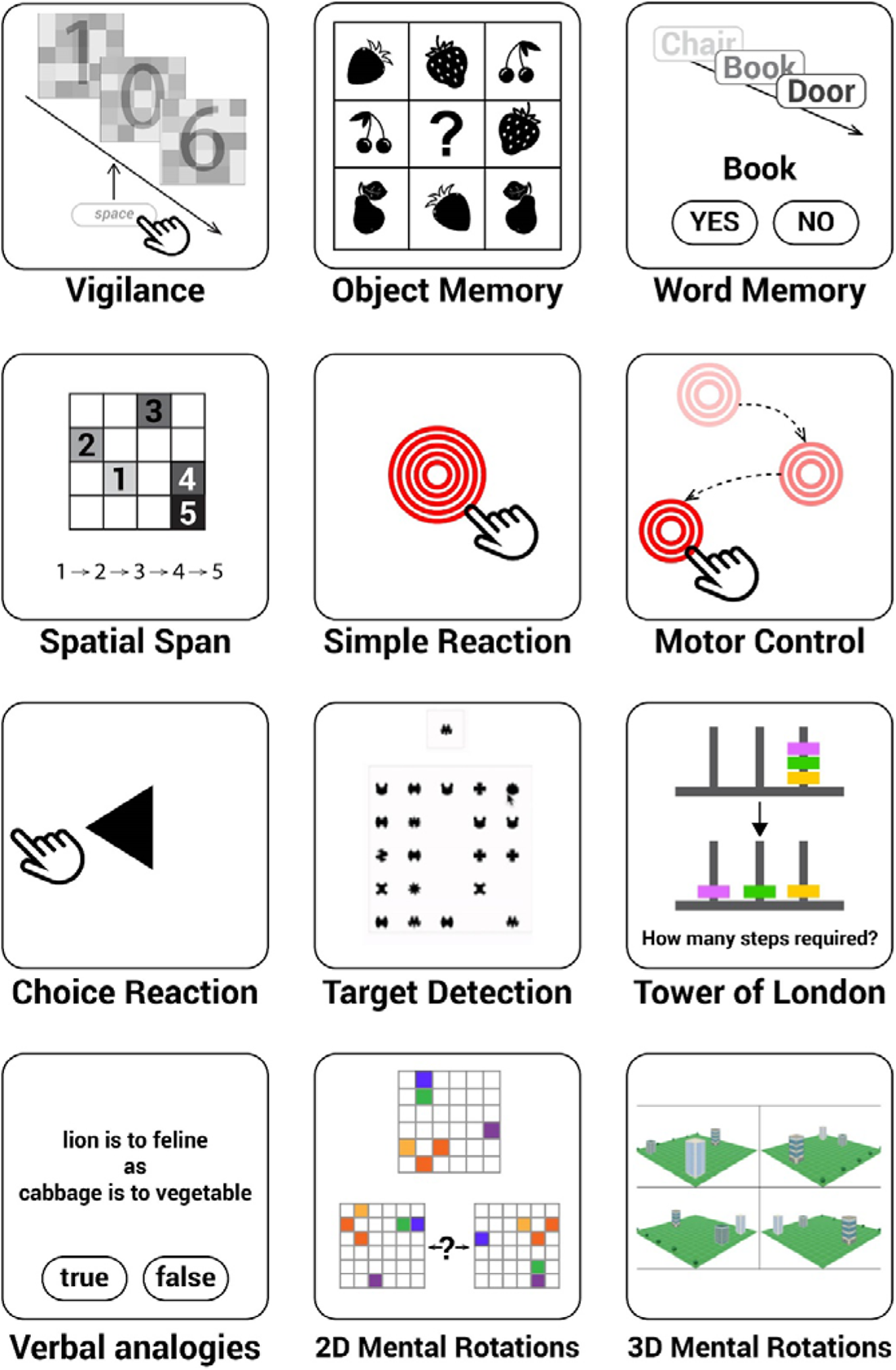
Twelve cognitive tasks measured distinct aspects of human cognition, memory, attention, motor control, planning and verbal reasoning abilities. The vigilance task was tested through the online experiment hosting server Pavlovia and conducted first. The rest of the 11 tasks were provided by Cognitron and ran in the following order: Motor control, Object memory (immediate), Word memory (immediate), Simple reaction task, Choice reaction time, 2D mental rotations, 3D mental rotations, Spatial span, Target detection, Tower of London, Verbal analogies, Object memory (delayed) and Word memory (delayed). The object memory and word memory tasks were both tested twice: once at the beginning (“immediate”) and again at the end of the experiment (“delayed”), with an interval of about 30 minutes. The delayed task was solely testing memory of the stimuli displayed in the first instance of the task so the memory probes were not displayed before the delayed task.

The experiment was implemented using PsychoPy v2021.1.2.^25^ To minimise the variance in latency caused by different browsers, all participants were required to use the Chrome browser on a desktop computer, although the operating system was not restricted. To minimise potential instability, the use of custom-written code was deliberately avoided and all functions used were as provided by PsychoPy. Only the questionnaires were implemented using in-house code.

#### Motivation and fatigue ratings with time on the task

After each minute during testing, participants were asked to report their level of fatigue (“How tired do you feel now?”) and motivation (“How motivated do you feel?”) using a visual analogue scale. Responses were registered by clicking on the appropriate position on each scale. After completing all ratings, a “confirm” button appeared at the bottom of the screen, allowing participants to validate their ratings and start the next block. To control the time between blocks and to reduce variance between participants, a 15-second countdown timer was displayed at the top of the screen, and the next block would begin automatically once the timer lapsed. There were seven participants who had missing ratings. The exclusion of these participants did not affect the rating results or behavioural data, and there were no group differences in age, gender or any questionnaire measures (*p*>0.05).

#### The session duration

The session of the questionnaires and vigilance test took controls 22.4 minutes (SD 6.7) on average and COVID survivors 23.8 minutes (SD 6.4). There was no statistical difference in the session length between two groups (t(135)=1.3, *p*=0.2). Although there was a relatively large variance in session duration amongst the participants, the duration of the main vigilance test was fixed at 11 minutes 15 seconds because each block was exactly 1 minute plus a fixed break of 15 seconds (a countdown was shown on the screen). The participants could take breaks during the questionnaire and the practice, the length of which were not recorded. Importantly, the vigilance decrement (see below) did not correlate with the session duration (Spearman rho=-0.06, *p*=0.5).

### Statistical analysis

#### Measuring of vigilance decrement

The accuracy of each minute was measured as an F1 score, the harmonic mean of precision and sensitivity, given by

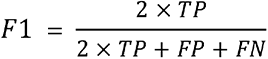

where TP is the true positive rate, FP is the false positive rate, and FN is the false negative rate. F1 score is a particularly suitable accuracy measure for this task because it emphasises true positives (responses to the rare target) and disregards true negatives (non-responses to non-targets). For RT, only trials with correct responses were considered. In each block, trials with outliers (two standard deviations away from mean) were excluded, representing 3.7% of trials overall.

To investigate the effect of group and time on minute-wise performance and the three ratings, all the values were z-scored across participants and mixed-effects generalised linear models (GLM) were conducted using the MATLAB function *fitglme* with Laplace approximation as the method for estimating model parameters. The models included fixed effects of group and time, and random effects of participants and their age.

The vigilance decrement for each individual was computed by subtracting the average accuracy over the first three minutes from the average over the last three minutes and then normalised by dividing the average over the first three blocks. The reason for using the average over the first three blocks as a performance baseline is because, in COVID-19 participants, the vigilance decrement happened relatively quickly, at the group level the accuracy was significantly lower than the first minute from the 4th minute (**Fig 3A**).

**Figure 3.**
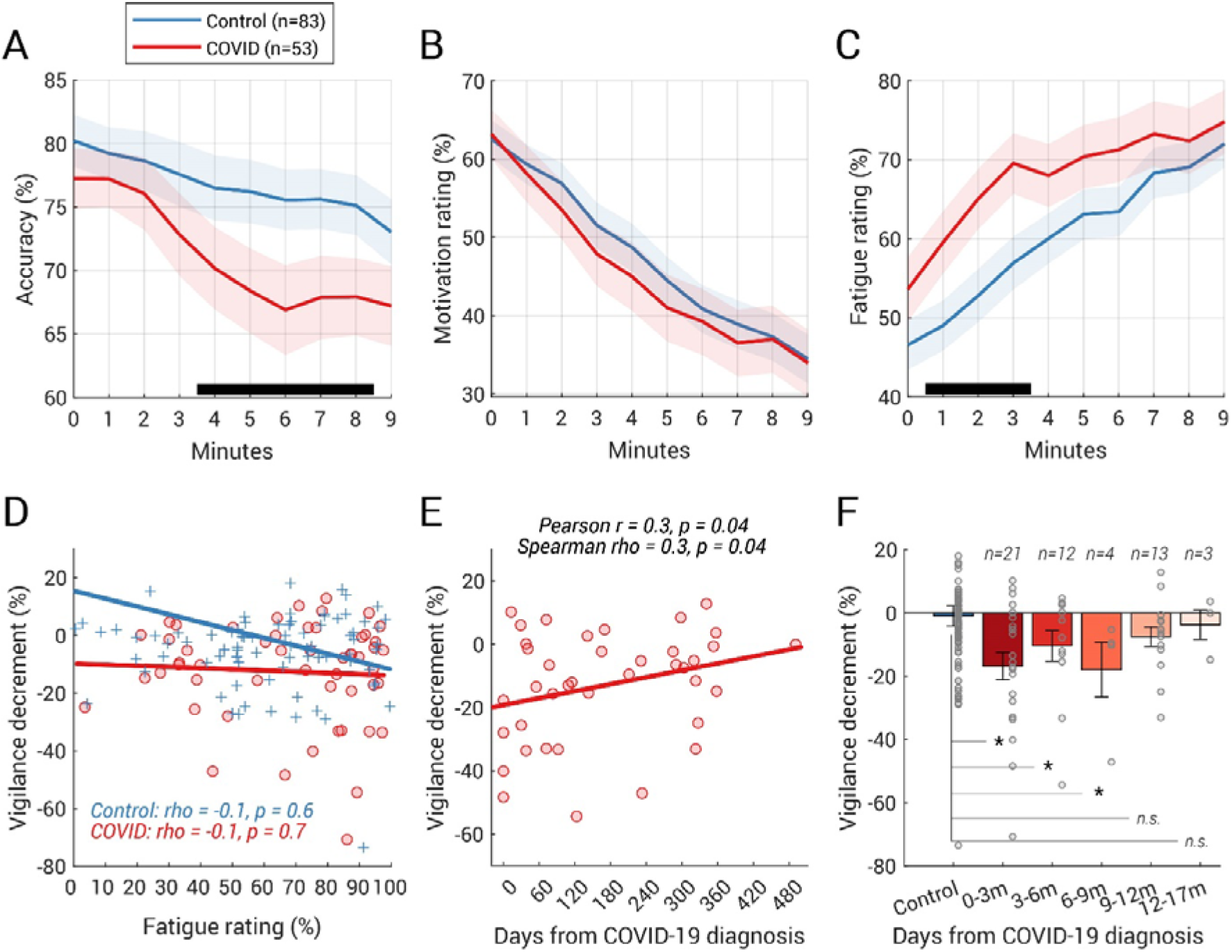
COVID group showed a larger and faster vigilance decline on the task. (A) Accuracy rate was computed for every minute (i.e., every block) of the vigilance test and plotted against the time. The y-value at t=0 corresponds to the accuracy rate over the one-minute-long practice block. The shaded area shows ±1 SEM and the black horizontal line at the bottom indicates time intervals where bootstrap statistics confirmed significant differences between the two groups (p<0.05, details see “Time-series analysis” in “Vigilance test”); the divergence was significant from the 4^th^ minute to the 8^th^ minute. (B) Group average of self-reported ratings of motivation against time (shaded area shows ±1 SEM). The rating at t=0 corresponds to the rating after the practice block. No group difference was found in the motivation rating over time. (C) COVID-19 survivors felt more tired from the beginning (shaded area shows ±1 SEM). (D) However, the fatigue rating (averaged over all 10 ratings) did not correlate with the size of the vigilance decrement in neither the COVID group nor the control group. The Spearman’s correlation coefficient and their two-tailed p values was shown for each group. (E) Vigilance decrement showed a significant correlation with the time from COVID-19 diagnosis. Both Spearman’s and Pearson’s correlation coefficients and their two-tailed p values are shown above the plot. (F) Participants who had COVID-19 within the last nine months displayed significantly larger vigilance decrements than the controls. The number of participants for each bin was labelled above each bar. Each grey dot represents individual data, and the error bar indicates one standard error. Group comparison performed by permutation test (with 10000 iterations). * = p<0.05, m (months).

#### Time-series analysis

To identify time intervals in which groups exhibit accuracy rate difference, a non-parametric bootstrap-based statistical analysis was used.^26^ In each iteration, N data sets (N=53 here; based on the number of participants in the COVID group) were selected with replacement from each group and a difference between means was computed. These time series were subjected to bootstrap re-sampling (10000 iterations). At each time point, differences were deemed significant if the proportion of bootstrap iterations that fell above or below zero was more than 95% (i.e., *p*<0.05). This analysis was applied to **Figs 3A, 3B**, and **3C**.

#### Group comparison

T-test statistics and Bayesian Factor (BF) are reported. All p values reported are two-tailed. When the sample sizes are not balanced, a bootstrap-based permutation test was used to confirm the reliability of the significance. For example, when comparing the vigilance decrement in participants who had COVID within two months (N=13) vs the control participants (N=73), in each iteration, N data sets (N=13 here; based on the number of participants who had COVID-19 within two months) were selected with replacement from each group and a difference between means was computed. This process was then repeated 10000 times. Group difference was deemed significant if the proportion of bootstrap interactions that fell above or below zero was more than 95% (i.e., *p*<0.05).

#### Correlation

To control for outlier effects, all reported bivariate and partial correlations were performed using the conservative Spearman’s rank correlation method. Additionally, we conducted partial correlations after controlling for the effect of the grit scale of personality.

### Cognitive test battery

On a subsequent occasion, a subset of participants also completed a sequence of eleven cognitive tasks to measure distinct aspects of human cognition, episodic memory, attention, motor control, planning, mental rotation and verbal reasoning abilities (**Fig 1**). These eleven tasks can be viewed at https://oxmh1.cognitron.co.uk. They were adapted from established behavioural paradigms in a manner that is sensitive to population variables of interest whilst being robust against the type of device that a person is tested on. The tasks were conceived, designed and programmed in HTML5 with JavaScript by A.H. and W.T., and hosted on a custom server system ‘Cognitron’ developed by P.J.H. on the Amazon EC2 platform.

This study was conducted independently to the vigilance task. All 155 participants were invited to attend this experiment, and 81 completed this part of the study between 6 and 20 May 2021, with 36 from the COVID group (mean time since COVID-19 diagnosis: 233.8 days (129.1), age 27.4(8.6), 14 females) and 44 from the control group (age 26.3(8.0), 17 females). No significant difference in age (t(80)=1.8, *p*=0.08, BF=1.1) or gender (χ^2^(1,N=31) = 0.0005, *p*=1.0), or any questionnaire-derived measures (all *p*>0.1) were observed in this subset of participants.

#### Object Episodic Memory

Participants were asked to memorise 20 images, all everyday objects, depicted in black and white. The images were presented sequentially in a random order, each for 2000 ms with an inter-image interval of 500 ms. Immediately after the presentation, participants’ memory of the 20 images was tested in 20 randomly ordered trials. Each trial required participants to select a previously displayed image from a set of eight images; incorrect images differed in the object itself, look or orientation (see the example in **Fig 2**’s *Object Memory*) in order to measure not only whether the correct target was identified, but also at higher precision the similarity of selected objects to the original target when errors were made. A delayed memory recall test was repeated at the end of the experiment, i.e., without additional sequential encoding of images; on average the time between the first and second test was 28.8 minutes. Population mean of the immediate accuracy was 60.1% (20.9) compared with the delayed accuracy of 57.6% (21.0), both well above chance level (12.5%). The memory decrement was computed by subtracting the accuracy in the immediate memory test from that in the delayed memory test for each individual.

#### Word Memory

This task involved memorising a sequence of English words (e.g.: Peas, Monkey, Dress, Aubergine, Mouse etc). Words could be randomly drawn from three categories: animals, vegetables or clothes. The words were presented sequentially in a pseudo-random order, each presented for 1000 ms and with an inter-image interval of 200 ms. Immediately post-presentation, word memory was tested with 24 words, including 50% non-targets, of which half were foils selected as being semantically similar to the targets (population mean of accuracy 90.9% (6.8)). After an average of 27.8 minutes later, participants were tested again to assess the episodic memory (mean accuracy (86.3% (9.1)). The memory decrement was computed by subtracting the accuracy in the immediate memory test from that in the delayed memory test for each individual.

#### Spatial Span

This task measured spatial short-term memory capacity. It was a variation of the Corsi Block Tapping Test^27, 28^ participants were presented with a 4×4 grid where each individual cell can light up sequentially. Participants were required to memorise the sequence and replicate it by clicking the appropriate squares in the order they lit up. The difficulty was incremented using a ratchet system, every time a sequence was recalled correctly, the length of the subsequent sequence was incremented by one. The test was terminated when three consecutive mistakes were made on a particular sequence length. The outcome measure was the maximum sequence length correctly recalled. Minimum level = 2, maximum level = 16, ISI = 0 ms, encoding time = 1500 ms. Mean sequence length successfully recalled was 6.6 (SD 1.3).

#### Simple Reaction Task

This task measured the basic cognitive process of perception and response execution. The participants were instructed to click on a red circle as soon as it appeared at the centre of the screen. They were presented with 60 stimuli, the ISI was jittered using a uniform random distribution between 0.5 s and 2 s. The dependent measure was the speed of response (population mean was 0.3 seconds (SD 0.1)).

#### Motor Control

This task was identical to the reaction time task above, except that the location of the red circle was different each time. Participants had to move their cursor to the target and respond as fast as possible. The participants were presented with 30 stimuli, the next stimulus was presented with a 0 ms delay after the response to the previous stimulus. Here, the population mean was 0.7 seconds (SD 0.2).

#### Choice Reaction Time

A black arrow either pointing left or right appeared on-screen, indicating the side of the screen the participant needed to click as fast as possible. The participants were presented with 60 stimuli with a 50% chance of each stimulus pointing left. The ISI was jittered using a uniform random distribution between 0.5 s and 2 s. Mean reaction time (RT) over button presses for both sides was computed (population mean was 0.5 seconds (SD 0.1)).

#### Target Detection

This spatio-visual attention task involved identifying and clicking on a target shape amongst a field of distractor shapes. The participant was presented with a target shape on the left of the screen and a probe area on the right side of the screen. After 3s, the probe area began to fill with shapes, the participant must identify and click the target shape while ignoring the distractor shapes. Shapes were added every 1s and a subset of the shapes in the probe area are removed every 1s. The trial ran for a total of 120 addition/removal cycles. The target shape was included in the added shapes pseudo randomly, at a frequency of 12 in 20 cycles. The outcome measure was the total number of target shapes clicked. Population mean was 60.2 (SD 10.4).

#### Tower of London

This task measured spatial planning and was a variant of the original Tower of London task.^29^ The participant was shown two sets of three prongs with coloured beads on them. The first set was the initial state and the second set was the target state. The task was to work out the lowest number of moves it would take to transition from the initial state to the target state, then input this number using an on-screen number pad. The test consisted of 10 trials of variable difficulty, scaled using the number of beads and the convolutedness, defined as the number of moves that must be made that do not place a bead in its final target position. The outcome measure was the total number of correct trials. Unlike in the original task^29^, the pegs were of equal height and the task was done mentally— the beads could not be moved. The total number of correct trials was recorded (population mean was 5.8 (SD 2.7)).

#### Verbal Analogies

This task examined semantic reasoning. Participants were presented with two written relationships that they must decide if they had the same type of association or not (e.g. “Lion is to feline as cabbage is to vegetable”). Participants indicated their decision by selecting the True or False buttons presented below the written analogies. Analogies were varied across semantic distance to modulate difficulty and association types switch throughout the sequence of trials. To obtain maximum points, participants must solve as many problems as possible within three minutes. For every correct response, the total score increased by one. For every incorrect response, the total score decreased by one. The outcome measure was the total score (population mean total point score was 19.9 (SD 12.9)).

#### 2D Mental Rotation

The 2D Mental rotation test measured the ability to spatially manipulate objects in the mind.^30^ In this version, we used 6×6 grids with various arrangements of coloured cells. Provided with a target grid and a set of four further grids, participants had to identify which of the four was the target but merely rotated by 90, 180 or 270° (incorrect grids had five incorrectly coloured cells). Participants were given three minutes to answer as many as possible, with correct trials being awarded with one point to their score (population mean was 34.5 (SD 8.5), technical issues beyond our control caused the loss of five participants’ records for this task).

#### 3D Mental Rotation

Akin to the 2D version above, participants were tasked with recognising a rotated version of the target from four options. In this 3D version, the grids were instead 3D scenes of buildings arranged upon a green surface. Again, the correct answer was identical to the target but from a different viewpoint (rotated by 90, 180 or 270°), while the others had their buildings in the wrong locations. Each participant was presented with 12 trials scored in the same manner as before (population mean was 4.1 (SD 6.2)).

#### The session duration

This session took controls 37.4 minutes (SD 13.6) on average and COVID survivors 35.3 minutes (SD 11.7). There was no statistical difference in the session length between two groups (t(94)=-0.8, *p*=0.4). The memory decrement (see below) did not correlate with the session duration (Spearman rho=0.1, *p*=0.4).

### Statistical analysis

For each cognitive function, a Principal Component Analysis (PCAs) was performed to derive a global index (PCA scores) quantifying its level across all relevant tasks used to assess it:

● Short-term memory score: PCA of full recall correct rate of *Object Memory*, full recall correct rate of *Word Memory*, and *Spatial Span* memory capacity.
● Executive and attentional function score: PCA of reaction times in *Simple Reaction Task, Motor Control, and Choice Reaction Time* and the overall score in *Target Detection*.
● Mental Rotation score: PCA of scores in *2D* and *3D Mental Rotation.* This quantifies each individual’s mental ability to spatially manipulate objects.

Group comparisons and correlations were performed as in *Vigilance*.

As the cognitive battery test was used as a broad-brush way of assessing a wide range of cognitive functions, Bonferroni correction was applied to the significant p value(s). Unadjusted estimates would be provided if stated, along with adjusted estimates reported in Results.

### Ethics statement

All participants gave electronic informed consent prior to the experiment. The study was approved by the local ethics committee and carried out in accordance with the relevant guidelines and regulations.

### Data availability

The data supporting the findings of this study are available from the corresponding author, upon reasonable request. A demo of the vigilance task is available at https://run.pavlovia.org/sijiazhao/vigilance_english_demo (please use the Chrome internet browser on a desktop computer). The experimental code is available at https://gitlab.pavlovia.org/sijiazhao/vigilance_english_demo.

## Results

COVID-19 survivors in this study were young, with many testing positive for COVID-19 several months before attending this study. Compared to the age-matched control group who did not report contracting COVID-19 before, there were no difference in gender or baseline group differences in a wide range of measures including fatigue/sleep abnormality (NFI), motivation (AMI), distractibility/forgetfulness (CFQ), mood (HADS) and personality (GRIT-S and BFI-S) (**Table 1**).

Compared with the controls, COVID-19 survivors showed significantly larger decline in performance in a 9-minute-long sustained visual attention task (**Fig 1** *Vigilance*). There was no significant group difference in accuracy over the baseline (i.e. the first three minutes, t(135)=-0.9, p=0.4, BF=3.8). However, COVID-19 participants showed a greater and more rapid decline in accuracy over time (significant interaction between group and time using a mixed-effects generalised linear model (GLM) (F(1,1346)=18.8, p<0.0001) along with main effects of time (F(1,1346)=15.1, p=0.0001) and group (F(1,1346)=8.1, p=0.005)). Comparing the minute-by-minute accuracy rate (**Fig 3A**), the group difference began to emerge by the start of the 4^th^ minute and ended after the 8^th^ minute.

On average, control participants’ accuracy dropped from 78.5% (SD 20.2, average accuracy over the first three minutes, see Table 2 for more details) to 75.4% (SD 20.9, average accuracy over the last three minutes), while COVID survivors started with a similar baseline at 75.5% (19.2), reducing to 67.8% (23.0) by the end of the ninth minute. Comparing the absolute change in performance over time, COVID-19 survivors showed a significantly larger decline (t(135)=-2.7, p=0.008, BF=5.0). Importantly, we attained a similar result after normalising the change by individual’s baseline performance (COVID -12.3% (17.4) vs Control -0.9% (29.1), t(135)=-2.6, p=0.01, BF=3.8) suggesting that this larger vigilance decrement amongst the COVID group was regardless of the individual’s baseline performance.

**Table 2.**
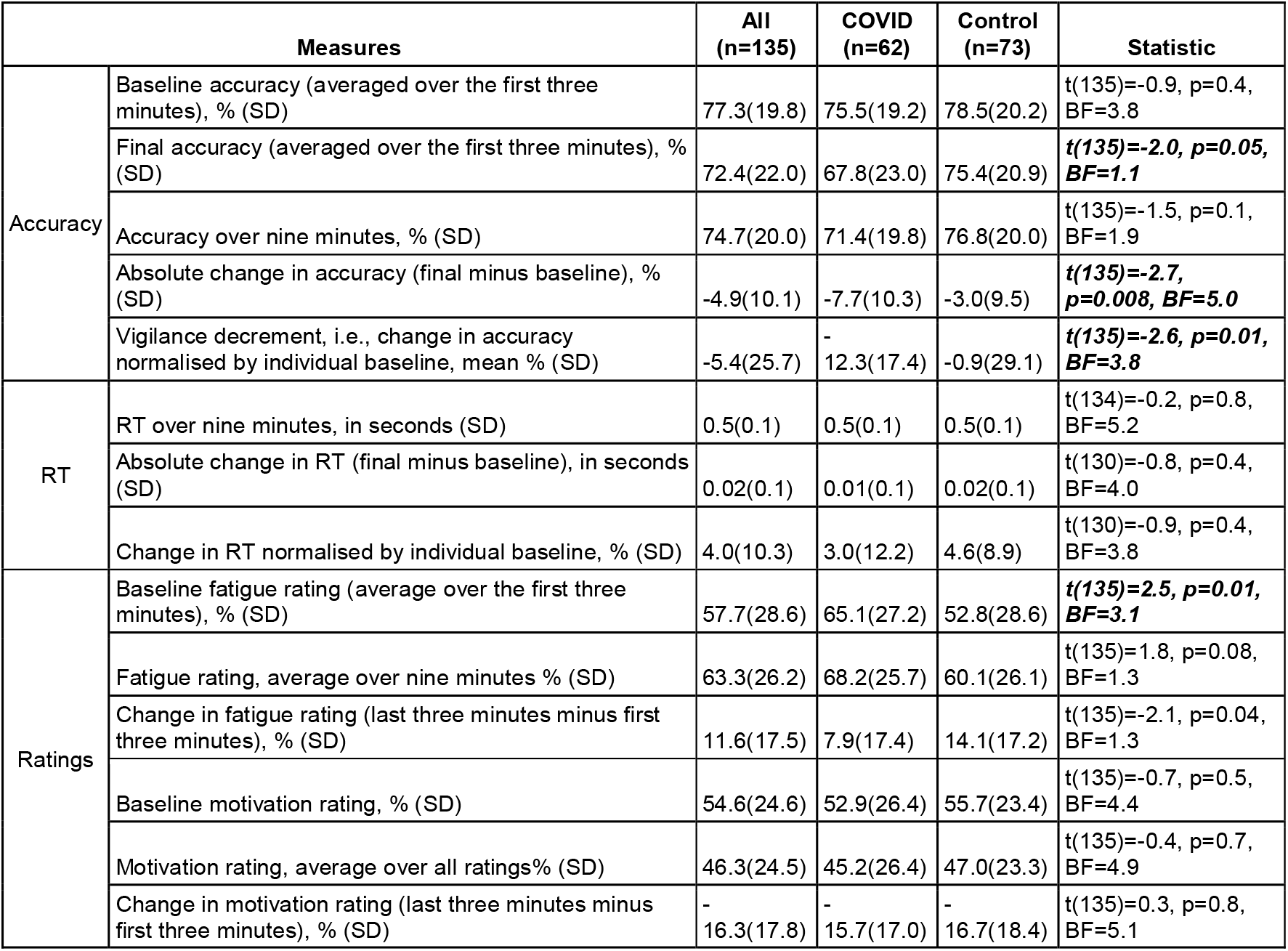
Results of Vigilance test. T-tests used to assess between-group differences, with Bayes Factor (BF) reported. The significant t-tests are highlighted in bold.

Across both groups, lower motivation (**Fig 3B**) and greater fatigue (**Fig 3C**) were reported as the experiment progressed (mixed effects GLM, main effect of time *p*<0.001). Crucially, the COVID group showed a significantly faster fatigue accumulation (interaction of group x time, F(1,1347)=8.8, *p*=0.003) and significantly larger fatigue rating over time (main effect of group, F(1,1347)=5.7, p=0.02). Although the COVID group had a statistically similar fatigue rating before the experiment started (t(131)=1.4, p=0.2, BF=2.3, COVID: 53.6%, SD 29.5, Control: 46.6%, SD 28.7), they started to report a significantly higher fatigue rating after completing the first minute of the test (**Fig 3C**), but these were not significant for motivation ratings (**Fig 3B**). Noticeably, the fatigue rating — neither the average over all 10 ratings, the baseline rating, nor the change in the rating in the first minute — correlated with the vigilance decrement (**Fig 3D**). This suggests that on an individual level, the vigilance decrement does not merely reflect the subjective feeling of being tired.

There was a significant positive correlation between individual vigilance decrement (normalised difference in average accuracy between first and last three minutes) and time from COVID diagnosis (**Fig 3E**, rho=0.3, *p*=0.04, two-tailed; Partial correlation after controlling age and personality of grit scale: rho=0.4, *p*=0.03, two-tailed). Furthermore, participants who had COVID-19 in the last nine months (**Fig 3F**) showed a significantly larger vigilance decrement than the control group (n=37, t(118) = -2.7, *p*=0.009, BF=4.7, bootstrap-based p=0.001), with this difference no longer apparent in participants who had COVID-19 more than nine months previously (n=16, t(97)= -0.8, *p*=0.4, BF=2.7, bootstrap-based *p*=0.2).

In order to obtain a more comprehensive examination of our COVID-19 survivors’ cognitive profiles, we subsequently invited all participants to complete a cognitive battery comprising 11 cognitive tests (all bar *Vigilance* in **Fig 2**) two months after the *Vigilance* test. 36 COVID-19 survivors had been diagnosed on average 233.8 days previously (129.1) and 45 age-matched controls attended the battery (**Table 2** for demographics).

Memory was a key cognitive function measured in this battery. In one of the memory tests— *Object Memory*—participants were shown 20 images of everyday objects to memorise. Specifically, participants needed to remember not only the object (e.g. a spoon), but also its look (e.g. a spoon with a long handle) as well as its orientation (e.g. handle pointing towards top-right). This provides us details about the preciseness of the memory recalled. One novel feature of this test is that *Object Memory* was tested twice, once immediately after presentation and again around 30 minutes later, which provides a measure of memory decrement over time.

On average, the controls achieved 59.7% accuracy in the immediate memory test. COVID- 19 survivors showed a similar short-term memory performance to the control group (t(78)=- 0.02, p=1.0, BF=4.3; **Fig 4A** and see **Table 2** for more details). However, a difference emerged in the later (delayed) test; while the controls displayed no memory decrement (no difference from zero, one sample t-test: t(43)=0.9, p=0.4, BF=5.8), COVID-19 survivors showed a significant memory decrement (t(35)=-4.1, p=0.0003, BF=107.5), which was larger than in controls by 9.2% (t(78)=3.3, p=0.001, BF=23.2; **Fig 4B**). This group difference would survive through Bonferroni correction (29 statistical tests were done for all 11 cognitive battery tests, see **Table 3** and **Table 4**, adjusted *p*=0.029). Like the vigilance decrement, the memory decrement is computed as a normalised change in performance (i.e., the difference between delayed and immediate memory test, divided by the immediate memory test). Importantly, this difference in episodic memory decrement was not due to the variance in memory maintenance duration, because there was no difference in the duration between the immediate and delayed memory tests across the two groups (COVID: 27.9 minutes (8.7), Control: 29.6 minutes (9.9), t(78)=-0.8, p=0.4, BF=3.2).

**Figure 4.**
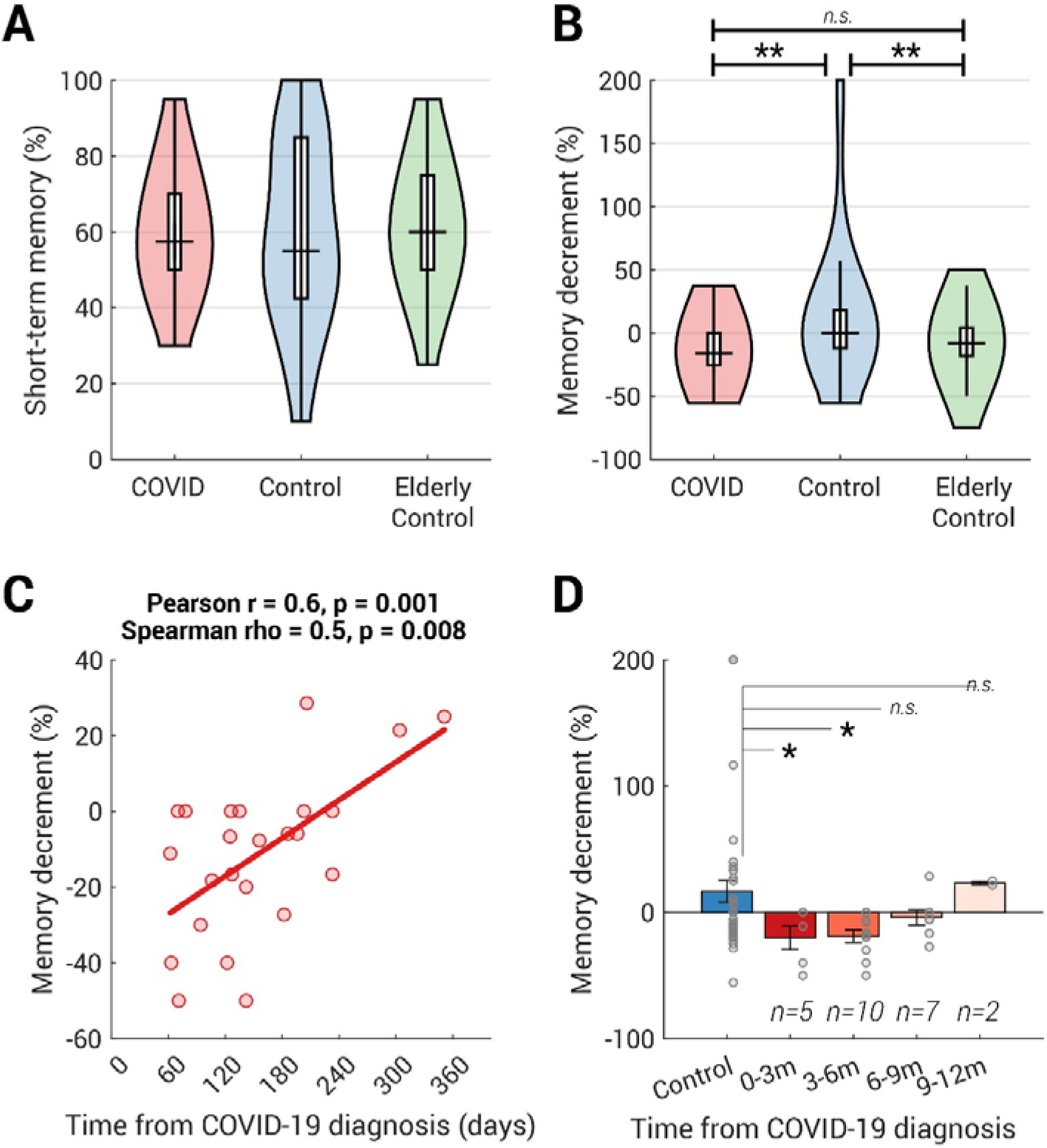
COVID group showed a mild episodic memory deficit compared with age- matched controls. (A) The distribution of the short-term memory, measured as the correct percent in the memory test immediately after viewing the sequence of objects, is plotted as a violin for COVID (n=36), Control (n=44) and Elderly Control (n=52, all above 50 years old, data collected separately). Group comparison performed by t-test. There were no statistical differences between groups in the short-term memory (COVID vs Control: t(78)=-0.02, p=1.0, BF=4.3; Control vs Elderly Control: t(94)=-0.4, p=0.7, BF=4.4; COVID vs Elderly Control: t(86)=-0.5, p=0.6, BF=4.0). (B) Approximately 30-minutes later, their memory was tested again. COVID and Elderly controls showed significantly larger memory decrements than the younger controls (COVID vs Control: t(78)=-3.0, p=0.004, BF=10.7; Control vs Elderly Control: t((94)=2.8, p=0.007, BF=6.0; COVID vs Elderly Control: t(86)=-1.4, p=0.2, BF=1.9). (C) In COVID-19 survivors who contracted COVID-19 within 1 year, the size of memory decrement was positively correlated with the time from COVID-19 diagnosis. Both Spearman’s and Pearson’s correlation coefficients and their two-tailed p values are shown above the plot. (D) Participants who had COVID-19 within the last six months showed significantly larger memory decrement than the age-matched controls. The number of participants for each bin was labelled above each bar. Each grey dot represents individual data, and the error bar indicates one standard error. Group comparison performed by permutation test (with 10000 iterations). * = p<0.05, ** = p<0.01, m (months).

**Table 3.** Demographics and the results of the Object Memory task in the battery test. The time from COVID-19 diagnosis was computed as the days between the date they attended this battery test and the self-reported date of their positive COVID-19 test. T- and *χ*^2^ tests used to assess between-group differences, with Bayes Factor (BF) reported. The significant t-tests are highlighted in bold.

| Measure |  |  | All<br>(n=80) | COVID<br>(n=36) | Control<br>(n=44) | Statistic |
| --- | --- | --- | --- | --- | --- | --- |
| Age, years, mean (SD) | | | 26.8(8.2) | 27.4(8.6) | 26.3(8.0) | $t(78)=0.6$, $p=0.6$, $BF=3.7$ |
| Gender, female, n (%) | | | 31(38.8) | 14(38.9) | 17(38.6) | $\chi^2(1, N=31) = 0.0$, $p=1.0$ |
| COVID-19 | Time from COVID-19 diagnosis, mean days (SD) |  |  | 233.8(129.1) |  |  |
|  | COVID-19 test type:<br>PCR / Lateral flow / Unknown / Not tested |  |  | 26/2/3/5 |  |  |
|  | Stayed at hospital overnight for COVID-19 treatment, yes (%) |  |  | 0(0.0) |  |  |
|  | Stayed at ICU for COVID-19 treatment, yes (%) |  |  | 0(0.0) |  |  |
| Object memory | Immediate memory test | Accuracy, mean % (SD) | 60.1(20.9) | 60.0(15.9) | 60.1(24.4) | $t(78)=-0.02$, $p=1.0$, $BF=4.3$ |
| | | RT in s, mean (SD) | 3.3(1.8) | 3.2(1.6) | 3.4(2.0) | $t(78)=-0.4$, $p=0.7$, $BF=4.0$ |
|  | Delayed memory test | Accuracy, mean % (SD) | 57.6(21.0) | 52.5(20.6) | 61.8(20.7) | **$t(78)=-2.0$, $p=0.05$, $BF=1.3$** |
| | | RT in s, mean (SD) | 2.3(0.8) | 2.2(0.6) | 2.4(1.0) | $t(78)=-1.3$, $p=0.2$, $BF=2.1$ |
|  | Memory decrement<br>(normalised by immediate accuracy) | Percentage change in accuracy, mean % (SD) | 3.0(46.8) | -13.6(21.4) | 16.6(56.9) | **$t(78)=-3.0$, $p=0.003$, $BF=10.7$** |
| | Time between immediate and delayed tests | Duration in minutes, mean (SD) | 28.8(9.4) | 27.9(8.7) | 29.6(9.9) | $t(78)=-0.8$, $p=0.4$, $BF=3.2$ |

**Table 4.** Results of the ten cognitive tasks in the battery test. Results from the same 80 participants reported in Table 3 are shown. T- and *χ*^2^ tests used to assess between-group differences, with Bayes Factor (BF) reported. Note that due to the technical issue, we lost five participants’ data for the 2D mental rotation task, thus resulting in a smaller degree of freedom in the t-test for that task.

| Measure |  |  | All<br>(n=80) | COVID<br>(n=36) | Control<br>(n=44) | Statistic |
| --- | --- | --- | --- | --- | --- | --- |
| Word memory | Immediate memory test | Accuracy, mean % (SD) | 90.9(6.8) | 91.1(7.1) | 90.7(6.7) | t(78)=0.2,<br>p=0.8, BF=4.2 |
|  |  | RT in s, mean (SD) | 1.0(0.3) | 1.0(0.3) | 1.0(0.2) | t(78)=0.2,<br>p=0.8, BF=4.2 |
|  | Delayed memory test | Accuracy, mean % (SD) | 86.3(9.1) | 86.3(8.6) | 86.2(9.5) | t(78)=0.1,<br>p=0.9, BF=4.3 |
|  |  | RT in s, mean (SD) | 0.9(0.2) | 0.8(0.2) | 0.9(0.2) | t(78)=-1.1,<br>p=0.3, BF=2.6 |
|  | Memory decrement<br>(normalised by immediate<br>accuracy) | Percentage change in<br>accuracy, mean % (SD) | -5.1(46.8) | -5.1(21.4) | -5.1(56.9) | t(78)=-0.04,<br>p=1.0, BF=4.3 |
|  | Time between immediate<br>and delayed tests | Duration in minutes,<br>mean (SD) | 27.8(8.4) | 26.7(6.8) | 28.8(9.5) | t(78)=-1.1,<br>p=0.3, BF=2.6 |
| Spatial short-term<br>memory capacity | Spatial span | n, mean (SD) | 6.6(1.3) | 6.4(1.4) | 6.8(1.3) | t(78)=-1.2,<br>p=0.2, BF=2.3 |
| Motor control | Simple reaction time | RT in s, mean (SD) | 0.3(0.1) | 0.3(0.1) | 0.3(0.1) | t(78)=-1.2,<br>p=0.2, BF=2.2 |
|  | Motor control | RT in s, mean (SD) | 0.7(0.2) | 0.7(0.2) | 0.7(0.2) | t(78)=0.5,<br>p=0.6, BF=3.9 |
|  | Choice reaction time | RT in s, mean (SD) | 0.5(0.1) | 0.5(0.1) | 0.5(0.1) | t(78)=-0.2,<br>p=0.9, BF=4.2 |
| Spatial visual<br>attention | Target detection | Score, mean (SD) | 60.2(10.4) | 60.4(11.0) | 60.1(10.0) | t(78)=0.1,<br>p=0.9, BF=4.3 |
| Spatial planning | Tower of London | Score, mean (SD) | 5.8(2.7) | 5.8(2.7) | 5.9(2.7) | t(78)=-0.2,<br>p=0.8, BF=4.2 |
| Semantic<br>reasoning | Verbal analogies | Score, mean (SD) | 19.9(12.9) | 18.3(11.2) | 21.2(14.2) | t(78)=-1.0,<br>p=0.3, BF=2.8 |
| Mental rotation | 2D mental rotation | Score, mean (SD) | 34.5(8.5) | 34.7(7.4) | 34.4(9.3) | t(73)=0.2,<br>p=0.9, BF=4.2 |
|  | 3D mental rotation | Score, mean (SD) | 4.1(6.2) | 4.3(6.1) | 4.0(6.3) | t(78)=0.3,<br>p=0.8, BF=4.2 |

Furthermore, the larger episodic memory decrement amongst COVID-19 survivors was driven by errors in which the wrong orientation was chosen for a correct item. In the immediate memory test, COVID-19 survivors had a 30.6% (13.3) false alarm rate where they chose the right object but wrong orientation, misbinding object identity with object orientation. This was not significantly different from controls (28.6 (18.3), t(80)=0.5, *p*=0.6, BF=3.8). However, this orientation-specific false alarm rate increased to 35.3% (15.7) 30 minutes later in the COVID group, which was significantly higher than the controls (27.3% (14.7), t(80)=2.4, *p*=0.02, BF=2.6). This difference suggests that the deficit in episodic memory in the COVID group might be associated with a deficit in binding information in memory.

In another memory test (**Fig 2** *Word Memory*), participants were instructed to memorise 24 simple English words. Although both COVID and control groups showed significant memory decrement (COVID: t(35)=-4.5, p=0.00008, BF=305.0; Control: t(43)=-4.4, p=0.00007, BF=314.5), they did not differ significantly from each other (t(78)=-0.04, p=1.0, BF=4.3). Because our participants reported having different first languages, we also ran a 2 (group: COVID vs control) x 2 (first language: English or non-English) ANOVA on the memory decrement. There were no main effects of first language (F(1,76)=1.1, *p*=0.3) or group (F(1,76)=0.1, *p*=0.7) and no language-group interaction (F(1,76)=0.1, *p*=0.7), indicating that this null effect was not due to differences in first language amongst participants. This null effect in word memory decrement might be due to the fact that the word memory task was much simpler than *Object Memory* which also had precision manipulation.

As part of a separate study, we ran the same memory tests on 52 healthy elderly participants (Elderly Control group: 53–82 years old, mean 67.4 (7.2), 30 females, no self-report COVID-19). Amongst controls (N=98, including all from Control and Elderly Control groups), there was a weak correlation between memory decrement and age (Pearson r=-0.2, *p*=0.03; Spearman’s rho=-0.2, *p*=0.07). Compared with young controls, elderly participants had a significantly larger memory decrement (mean (SD): -7.1 (21.8), t((94)=2.8, *p*=0.007, BF=6.0, **Fig 4B**), but importantly elderly participants’ episodic memory decrement was not statistically different from COVID-19 survivors (t(86)=-1.4, p=0.2, BF=1.9, **Fig 4B**), indicating that COVID-19 survivors performed as if they were older.

However, the elderly controls spent a longer time completing other tasks between the immediate and delayed memory tests (mean 39.2 (6.1) minutes) than the COVID group (t(86)=-7.2, *p*<10^9^, BF>10^7^). The requirement to maintain memory for a longer period was associated with greater memory decrements (partial correlation of time between memory tests and memory decrement amongst young and elderly controls after controlling the effect of age: Pearson r=0.2, *p*=0.03; Spearman rho=0.05, *p*=0.6). Therefore, we regressed out the effect of the memory maintenance time from each individual’s memory decrement, but the pattern remained unchanged.

Amongst survivors who contracted COVID-19 within the last year, the size of memory decrement after controlling for the effect of age was weakly but significantly correlated with the time from diagnosis (**Fig 4C**, Pearson r=0.6, *p*=0.001; Spearman rho=0.5, *p*=0.008), suggesting that people who had COVID-19 more recently tended to forget more over the 30- minute interval. This significant memory decrement could be observed up to 6 months (N=15, t-test: t(57) = -2.4, *p*=0.02; **Fig 4D**). These analyses provide preliminary evidence that COVID-associated reductions in sustained attention and episodic memory may persist for months, but may normalise subsequently, although the findings have to be taken with caution given the sample size.

Do these cognitive differences relate to the symptom severity experienced during COVID-19 illness or the post-illness long-COVID? In a follow-up survey, we asked the participants for their experience during and after their COVID-19 illness (**Fig 5A** **and** **Fig 5C**). The questions were modified from *Office for National Statistics – Coronavirus (COVID-19) Infection Survey*; for example, the question about the COVID-19 symptom severity was “Do any of the COVID- 19 symptoms reduce your ability to carry out day-to-day activities?” with four options: Yes, a lot/Yes, a little/Not at all/No symptom, which in turn corresponds to Severe/Moderate/Mild/Asymptomatic. To assess the relation between the post-COVID cognitive decrements observed here and the COVID-19/long-COVID severity, a linear mixed-effect model (LMM) with COVID-19 severity level and long-COVID severity level as fixed effects, and participant as a random effect was applied. Amongst the COVID-19 survivors who had any COVID-19 or long-COVID symptoms, larger vigilance and memory decrements were associated with more severe COVID-19 symptoms (**Fig 5B**, main effect on vigilance decrement: F(1,8)=6.8, *p*=0.03; main effect on memory decrement: F(1,6)=15.3, *p*=0.008) but not with long-COVID symptoms (**Fig 5D**, main effect on vigilance decrement: F(1,8)=4.5, *p*=0.07; main effect on memory decrement: F(1,6)=0.09, *p*=0.8). However, two caveats require attention: First, the positive relation between the COVID-19 symptom severity and the cognitive decrements must be taken with caution because it disappears if taking asymptotic participants into account. Secondly, the null effect of long-COVID severity on the cognitive decrements might be specific to the present study as most of COVID-19 survivors in the present study did not have any long-COVID symptoms. Nevertheless, further confirmation of these relations is out of scope of the present study and should be addressed in patient studies amongst COVID-19 inpatients and/or long-COVID patients.

**Figure 5.**
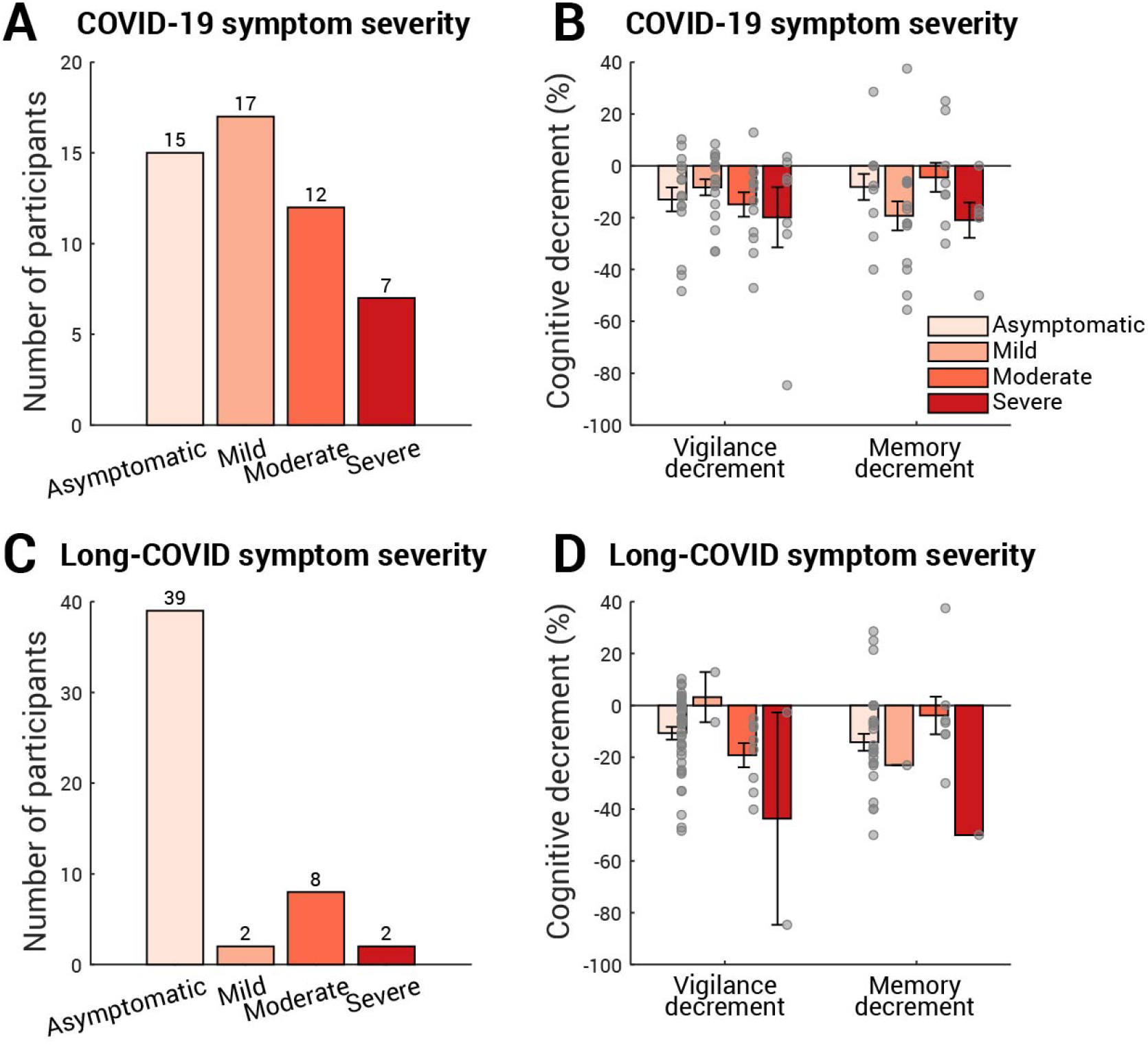
Cognitive decrements sorted by COVID-19 symptom and long-COVID symptom severity. 51 out of 64 COVID-19 survivors (including the three participants who stayed hospital overnight for COVID-19) reported their COVID-19 symptom severity (A) and long-COVID symptom severity (C). In both (A) and (C), the number of participants for each severity level is labelled above corresponding bar. (B) The vigilance (left) and memory (right) decrements binned by COVID-19 symptom severity. Each grey dot represents individual data, and the error bar indicates one standard error. A linear mixed-effect model (LMM) with participant as a random effect showed that COVID-19 severity level had main effect on vigilance decrement (F(1,8)=6.8, p=0.03) and main effect on memory decmrenet (F(1,6)=15.3, p=0.008). Similarly, (D) shows the cognitive decrements for each long-COVID symptom severity level. An LMM with participant as a random effect showed that long- COVID symptoms had no effect on vigilance decrement (F(1,8)=4.5, p=0.07) or memory decrement (F(1,6)=0.09, p=0.8).

Although the correlation between cognitive decrement and time since COVID-19 diagnosis (**Fig 3E** and **Fig 4D**) provides strong evidence linking cognitive differences observed in the present with COVID-19 infection, we additionally considered additional non-infectious factors. First, we ruled out basic factors including age (**Fig 6A**, t(134)=-0.6, *p*=0.5), gender (**Fig 6B**, *Χ*^2^ (1,N=54)=0.5, *p*=0.5), first language (**Fig 6C**, *χ*^2^ (1,N=50)=0.03, *p*=0.9), country of current residence (**Fig 6D**, *χ*^2^(1,N=36)=0.2, *p*=0.7), and ethnicity (**Fig 6E**, all categories *p*>0.1), as none showed any difference between the two groups. Secondly, we pondered whether the more significant decline in attention and memory of the COVID group could be attributed to a lower socioeconomic status (SES). This is highly possible: a recent study^31^ suggested strong associations between low SES and high probability of COVID-19 infection, along with higher infection fatality rate. Meanwhile, lower SES has established negative pressures on cognition including attention and memory (see review^32^). To address this concern, all participants received a follow-up survey covering a wide range of demographic and socioeconomic measures, encompassing education, income, occupation, work sector as well as subjective SES measured by the MacArthur Scale of Subjective Social Status^33^. 117 responded (75% of 155 participants, COVID N=51, Control N=66), with no statistical difference in education level (**Fig 6F**), annual income (**Fig 6G**) or employment status (**Fig 6H**). In fact, the COVID group reported a slightly higher subjective SES (**Fig 6I**, COVID subjective SES = 6.1(1.2), Control subjective SES = 5.6(1.5), COVID vs Control: t(96)=2.5, *p*=0.02). Moreover, we found no difference in the proportion of essential workers amongst the groups (**Fig 6J, *χ***^2^(1,N=51)=0.8, *p*=0.4) or the method of the commute during the pandemic (**Fig 7A**, all categories *p*>0.1). These suggest that SES cannot fully explain the cognitive difference between COVID and control groups in the present study. Another potential confound is testing experience; participants from the COVID group might, by chance, have less experience of cognitive testing than the controls. This hypothesis can be tested by comparing the number of studies that the participants attended on Prolific. In contrast, the COVID group had greater experience of online experiments (t(134)=4.4, *p*<10^-4^). The average number of studies attended was 248 studies (SD 358) in the COVID group and 67 (SD 82) in the control group. Moreover, the number of attended studies does not correlate with vigilance decrement (Spearman rho=-0.09, *p*=0.3) or memory decrement (Spearman rho=-0.05, *p*=0.7). These confirm that cognitive differences observed in the present study were not caused by the effect of test familiarity. Although it is unclear if smoking history could affect the cognitive ability, we also checked for this. Reassuringly, there was no difference in the proportion of past smokers (**Fig 7B**, *χ*^2^(1,N=29)=0.9, *p*=0.3) or present smokers (**Fig 7C**, *χ*^2^(1,N=14)=0.9, *p*=0.4) between the two groups. Finally, we asked for COVID-19 vaccination history: unsurprisingly, there was a significantly higher vaccination rate amongst controls than COVID-19 survivors (**Fig 7D**, *χ*^2^(1,N=15)=8.6, *p*=0.003). On average, the days from the last dose of vaccination to the date of attending the test was 63.7 (SD 37.9, minimum = 12, maximum = 139) and was not correlated with the vigilance decrement (Spearman rho=0.1, *p*=0.6). Taking all the evidence together, the results suggest that the cognitive differences in attention and memory observed here seem to be strongly associated with COVID-19 infection, rather than an outcome of a single demographic or socioeconomic metric.

**Figure 6.**
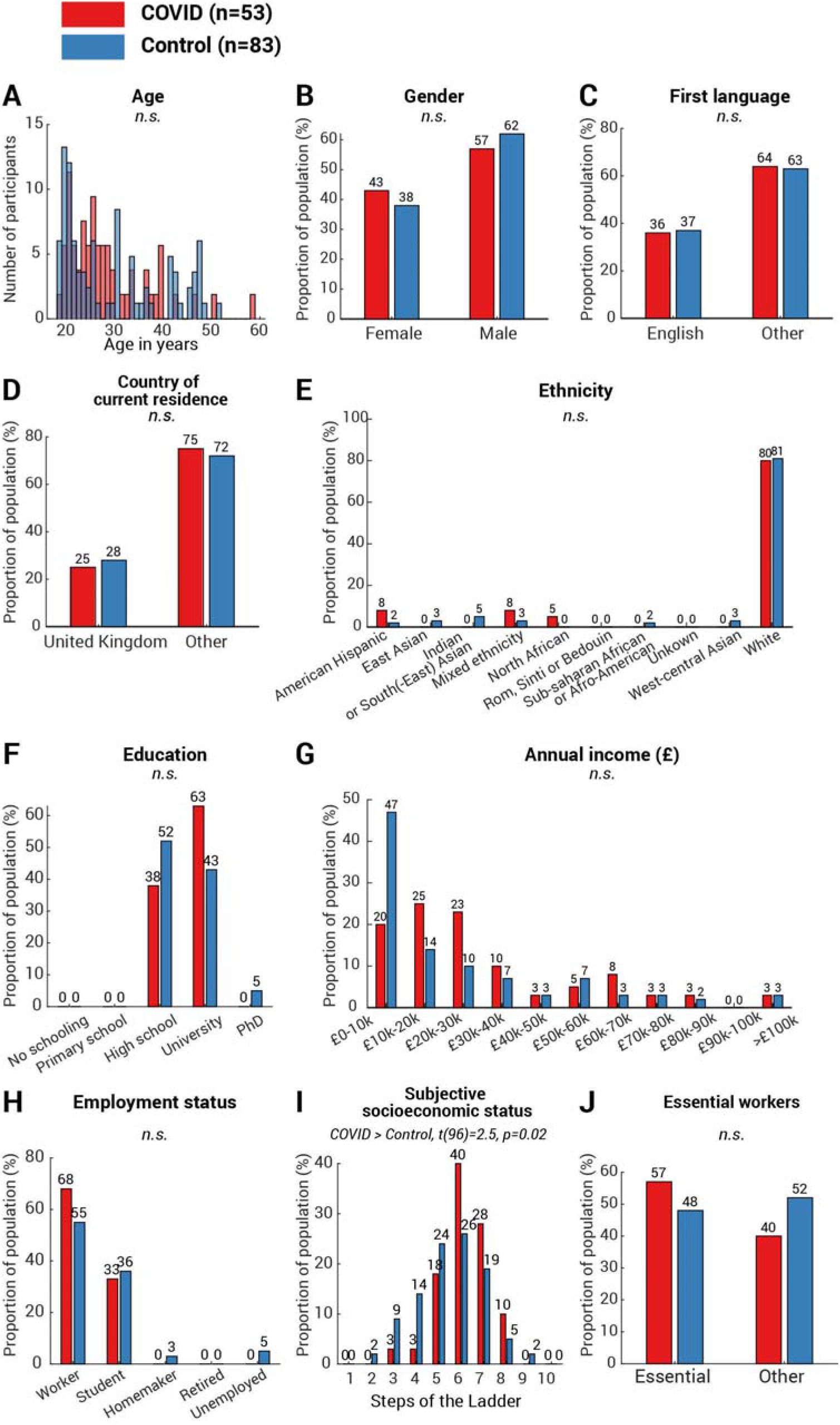
Demographics and socioeconomics profile of the participants. T-test was used to assess between-group difference in age (A) and subjective socioeconomic status (I). For the measures with binary outcomes, including gender (B), first language (C), country of current residence (D) and essential workers (J), *χ*^2^ test was used to assess between-group differences. Their p values were unadjusted for multiple comparison. For the measures with multiple categories—ethnicity (E), education (F), annual income (G) and employment status (H), *χ*^2^ test was run for each category of each measure, and p values were adjusted using the Bonferroni method (i.e., multiplying the number of categories in that measure). Amongst all measures, only one measure showed significant difference: The COVID group showed a significantly higher subjective socioeconomic status (h, t(96)=2.5, p=0.02). No difference was found in other measures and annotated as n.s. (not significant).

**Figure 7.**
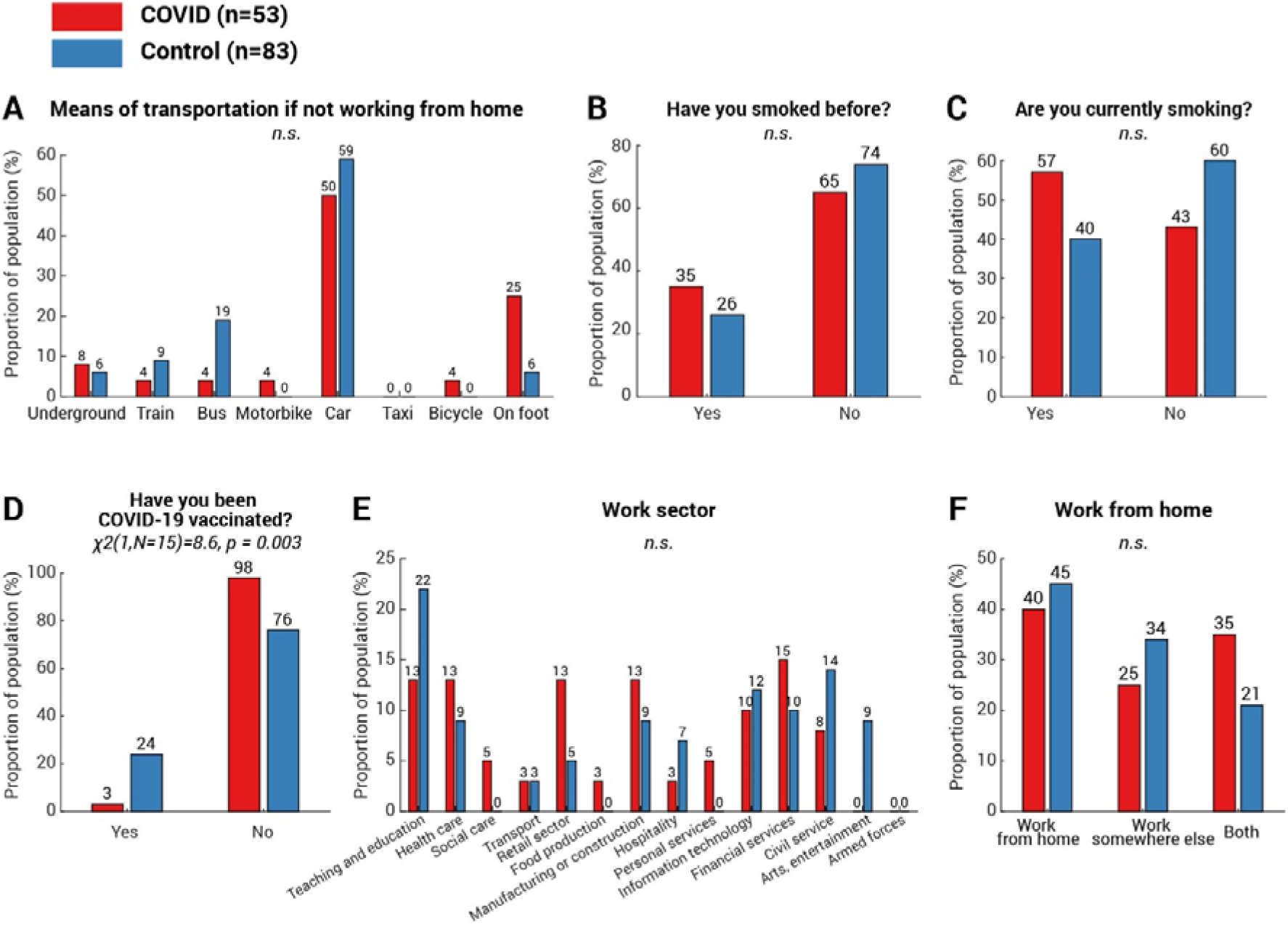
Work, smoking history and vaccination status of the participants. For the measures with multiple categories— transport means used to commute (A), work sector (E) and work from home status (F), *χ*^2^ test was run for each category of each measure, and p values were adjusted using the Bonferroni method (i.e., multiplying the number of categories in that measure). For the measures with binary outcomes, including smoking history (B and C) and COVID-19 vaccination history (D), *χ*^2^ test was used to assess between-group differences. Amongst all measures, only one measure showed significant difference: The COVID group showed a lower rate of COVID-19 vaccination (D, *χ*^2^(1,N=15)=8.6, p=0.003). No difference was found in other measures and annotated as n.s. (not significant).

Encouragingly, apart from these two cognitive differences, COVID-19 survivors did not show any significant difference from the age-matched controls in a wide range of cognitive capabilities, including short-term memory (*Object Memory, Word Memory, Spatial Span*), the response speed (*Simple Reaction Task, Motor Control, Choice Reaction Time*), spatial-visual attention (*Target Detection*), spatial planning (*Tower of London*), semantic reasoning (*Verbal Analogies*) and mental rotations (2D or 3D) (see **Table 3** and **Table 4** for details). Similarly, comparing PCA scores between COVID-19 survivors and controls, no significant difference was found in the short-term memory (t(78)=-1.2, p=0.2, BF=2.2), the executive function (t(78)=0.3, p=0.7, BF=4.1) or the mental rotation ability (t(73)=0.2, p=0.8, BF=4.2), suggesting that most of the key cognitive functions were normal.

## Discussion

In the present study, we examined a wide range of cognitive abilities in COVID-19 survivors and age-matched controls. The COVID group did not require hospitalisation and had not sought medical help for long COVID symptoms after recovery. The good news is that COVID-19 survivors performed well in most cognitive abilities tested, including working memory, executive function, planning and mental rotation. However, even though their questionnaire-derived measures (fatigue, forgetfulness, motivation, sleep abnormality, depression and anxiety levels) were no different from age-matched controls (**Table 1**), they showed a significantly larger vigilance decrement along with faster fatigue build-up over the course of a 9-minute-long attentionally demanding task (**Figs 3A** and **3C**). They also had significantly worse episodic memory decrement over time, comparable to a healthy, elderly person in their 60s (**Fig 4B**). Notably, both deficits scaled with the time from COVID-19 diagnosis suggesting a strong relation with COVID-19 itself (**Figs 3E** and **4D**).

To our knowledge, this is the first report describing deficits in sustained attention and episodic memory amongst mildly-affected COVID-19 survivors long after the acute illness, in people who were not complaining of long-COVID symptoms (cf. Zhou et al.^9^ reporting sustained attention impairments in recently-recovered patients). Our findings are consistent with the most prevalent complaints concerning post-COVID cognitive issues, including poor concentration and/or impaired memory in 18-50% of patients post-recovery.^8,34, 35^ However, in contrast with previous reports of long-COVID patients^6–8, 12, 13, 16^, here we found no difference in short-term attention (performance over the first few minutes of a vigilance test; overall performance in tasks measuring executive function and response speed, **Table 4**) or working memory (performance in the immediate object or word memory tests, **Table 3**, **Table 4** and **Fig 4A**).

In the present study, COVID-19 survivors began with apparently normal behavioural performance followed by a gradual decline away from age-matched controls, suggesting reduced ability to attentively track and maintain information *over time*. The inconsistency with previous reports might be due to the fact that patients featured in those studies had severe COVID-19 symptoms^3, 4, 6, 12^, clinically significant cognitive impairment^16^, or at least reported persistent cognitive symptoms^7, 8, 13^, while our participants were mostly non-hospitalised and devoid of self-reported abnormality.

The mechanisms underlying these cognitive deficits as yet remains unclear. Although a direct effect of virus persisting in the brain cannot be excluded, the evidence from post-mortem studies suggests there is very little presence of virus within the brain in COVID-19 patients.^36^ Rather, there might be indirect effects of the virus on cognitive function mediated via a range of possible mechanisms, including immunological and microvascular changes (see review^37^). One investigation of COVID-19 survivors demonstrated that the most severely cognitively affected patients demonstrated a degree of cognitive impairment accompanied by hypometabolism in the frontoparietal regions.^10^ These brain regions are implicated in sustained attention^38^ as well as in episodic memory.^39–41^ Reassuringly, the follow-up study of Hosp and colleagues^10^ showed slow but evident improvement after six months.^11^ This is in line with the mildly-affected individuals reported here: both vigilance and episodic memory decrements gradually resolved over time (**Figs 3E** and **4C**). Episodic memory returned to normal levels after six months (**Fig 4D**) and those who had COVID-19 over nine months ago did not exhibit the vigilance decrement (**Fig 3F**).

Unlike other survey-based reports focusing on self-reported long-COVID symptoms^35, 42–44^, COVID-19 survivors in the present study did neither indicated any sign of higher fatigue, forgetfulness, apathy, anxiety, depression or sleep abnormality (**Table 1**), nor felt any more tired than their age-matched controls over the time course of the vigilance test, suggesting a dissociation between self-report symptoms and objectively measured deficits. Our findings highlight that cognitive reductions are not limited to patients who had prolonged neurological manifestations after recovery^7, 8^, but might exist more widely in a sub-clinical form amongst COVID-19 survivors who would not consider themselves requiring any post-COVID treatment.

At the outset of the pandemic, Hampshire et al.^17^ conducted a large-scale online test involving over 13,000 people with suspected or biologically confirmed COVID-19 circa two months. That study shared a subset of tasks with our investigation, covering a wide range of cognitive functions including semantic problem solving, visual spatial attention, speed of response and working memory. All functions showed some degree of cognitive deficit amongst people who had contracted COVID-19, scaling with respiratory symptom severity. In the present study, however, we did not find any group differences in these cognitive domains (**Table 2**). This difference is likely to be attributable to the relative mildness of the symptoms experienced by our COVID-19 survivors, combined with the length of time since infection (some over nine months ago). This suggests that these functional deficits might not be obvious in milder COVID-19 patients, with recovery expected within months.

There are some limitations to our study. Although the majority of participants from our COVID group reported a PCR-confirmed COVID-19 positive result with none reporting the need for post-COVID treatment, our study was limited by our reliance on self-reports of positive/negative COVID-19 tests and timing of diagnosis, which might increase or decrease our estimate of the prevalence and duration of COVID-associated cognitive deficits. Our study is also constrained by a relatively small sample size (N=136) with under- representation of the over 70s, thus any generalization should be taken carefully.

Overall, the findings here show that COVID-19 survivors showed a significant reduction in their ability to sustain attention on a demanding task up to nine months after COVID-19 infection, along with mild, but significantly worse, episodic memory for up to six months. Just as the acute illness of COVID-19 demonstrates a wide severity spectrum from asymptomatic to fatal forms^45^, our findings show that post-COVID cognitive deficits too can also manifest a wide severity spectrum. They highlight a pressing need to measure cognitive performance objectively in order to better understand how the brain is affected by COVID-19.

## Acknowledgements

The authors are grateful to Stephen Walsh for his involvement in the early stage of the project.

